# Trajectories of menstrual symptoms and blood pressure in midlife: A prospective cohort study on Australian women

**DOI:** 10.1101/2024.02.18.24303010

**Authors:** Gita D Mishra, Chuyao Jin, Hsiu-Wen Chan, Jenny Doust

## Abstract

**Aim:** To investigate the associations between the trajectories of menstrual symptoms over 18 years and blood pressure in midlife using longitudinal data.

**Methods:** Using data from the Menarche-to-PreMenopause (M-PreM) study, sampled from the Australian Longitudinal Study on Women’s Health, 494 participants were followed from their early 20s to mid-40s. Distinct trajectories of heavy menstrual bleeding, irregular periods, and severe period pain were identified by group-based trajectory modelling. The associations between menstrual symptom trajectories and blood pressure were examined by linear regression models.

**Results:** We identified three distinct heavy menstrual bleeding trajectories and two for irregular periods and severe period pain. After accounting for blood pressure monitors and socio-demographic factors, women in the chronic heavy menstrual bleeding group and late onset heavy menstrual bleeding group exhibited higher systolic (SBP) and diastolic (DBP) blood pressure compared to the normative group. The late onset irregular periods group also had higher DBP than the normative group. When we further adjusted for lifestyle factors, body mass index, and waist-to-hip ratio, the associations were attenuated (chronic heavy bleeding: SBP [B=3.0; 95% CI: −1.1, 7.0] and DBP [B=2.5; 95% CI: −0.3, 5.2]; late onset heavy menstrual bleeding: SBP [B=3.4; 95% CI: 0.3, 6.5] and DBP [B=3.1; 95% CI: 1.0, 5.2]). We found no associations between severe period pain and blood pressure.

**Conclusions:** The trajectories of heavy menstrual bleeding and irregular periods during the reproductive span were associated with blood pressure in midlife. This suggests that tracking menstrual symptoms in women could help predict their midlife blood pressure.

## Introduction

Up to 91% of women of reproductive age report experiencing menstrual symptoms, including painful menstruation (dysmenorrhoea), heavy or prolonged menstrual bleeding (menorrhagia), or irregular menstruation ^1–3^. The prevalence of each symptom is reported to vary, which may be due to a variety of lifestyle behaviours (e.g., body weight, smoking, physical activity, sleep) ^4, 5^, genetic factors (e.g., family history of menstrual disorders) ^6^, and reproductive history (e.g., history of preterm birth, age at menarche) ^1, 7^. Menstrual symptoms can dramatically affect quality of life and reproductive outcomes ^5, 7, 8^. They may also affect cardiovascular health in the long run, including blood pressure, through processes including increased arterial stiffness, an overactive renin-angiotensin-aldosterone system (RAAS), oxidative stress and activation of inflammatory pathways ^9–12^.

Studies investigating the relationship between menstrual symptoms and blood pressure tend to be cross-sectional, reporting changes to blood pressure and hormones or other factors that regulate plasma volume at different phases during the menstrual cycle in women and are also typically restricted to cohorts of women who experience pre-menstrual syndrome ^11, 13, 14^. To date, there have only been two studies that have assessed the prevalence of menstrual symptoms across the reproductive life course, with one reporting on the relationship between menstrual symptoms and hypertension. Chung et al. (2021) reported not only that heavy menstrual bleeding increased the incidence of hypertension by 53% but that the relationship may be bidirectional, with hypertension also increasing the incidence of heavy menstrual bleeding by 23% and irregular periods by 42% ^1^. Menstrual symptoms may change across the reproductive lifespan and different menstrual symptom trajectories affect up to 80% of women ^15^. To date, no studies have captured how the experience of chronic menstrual symptoms or the emergence of symptoms at different time points throughout the reproductive lifespan may affect blood pressure.

Therefore, this study used the Australian Longitudinal Study on Women’s Health (ALSWH) dataset to investigate the relationship between the trajectory of specific menstrual symptoms, including heavy menstrual bleeding, irregular periods and severe period pain, with changes to mean systolic (SBP) and diastolic blood pressure (DBP) across the course of 18 years.

## Methods

### Study population

Subjects were from the Menarche-to-PreMenopause (M-PreM) study conducted from June 2019 to June 2021, which aims to elucidate the associations of reproductive factors with cardiometabolic markers, respiratory conditions, and cognitive and functional health before women reach middle age^16^. Participants in the M-PreM study were from the Australian Longitudinal Study on Women’s Health (ALSWH), a national, prospective study following three cohorts of women (born in 1921-26, 1945-51, 1973-78) randomly selected from the Australia’s universal health insurance scheme (Medicare) since 1996. Questionnaires were sent to the women approximately every three years to collect information including their health status, lifestyle factors, and health service use. More details of ALSWH have been reported previously^17, 18^. Invitations to participate in the M-PreM study were sent to all women who were in ALSWH 1973-78 cohort and completed Survey 8 when they were 40-45 years old (2018, N=7121). Among 4584 women who expressed interest to participate, 499 of them finished clinical assessment and 779 did self-assessment. Women who completed clinical assessment and had blood pressure measurements were included in the analysis (N=494, Figure 1). Compared with women not included in the analysis, those included were younger, were more likely to live in major cities, had higher education and physical activity level, and were less likely to be current smokers. They were comparable regarding the use of oral contraceptive pill and the prevalence of menstrual symptoms (Supplemental Table 1).

**Figure 1.**
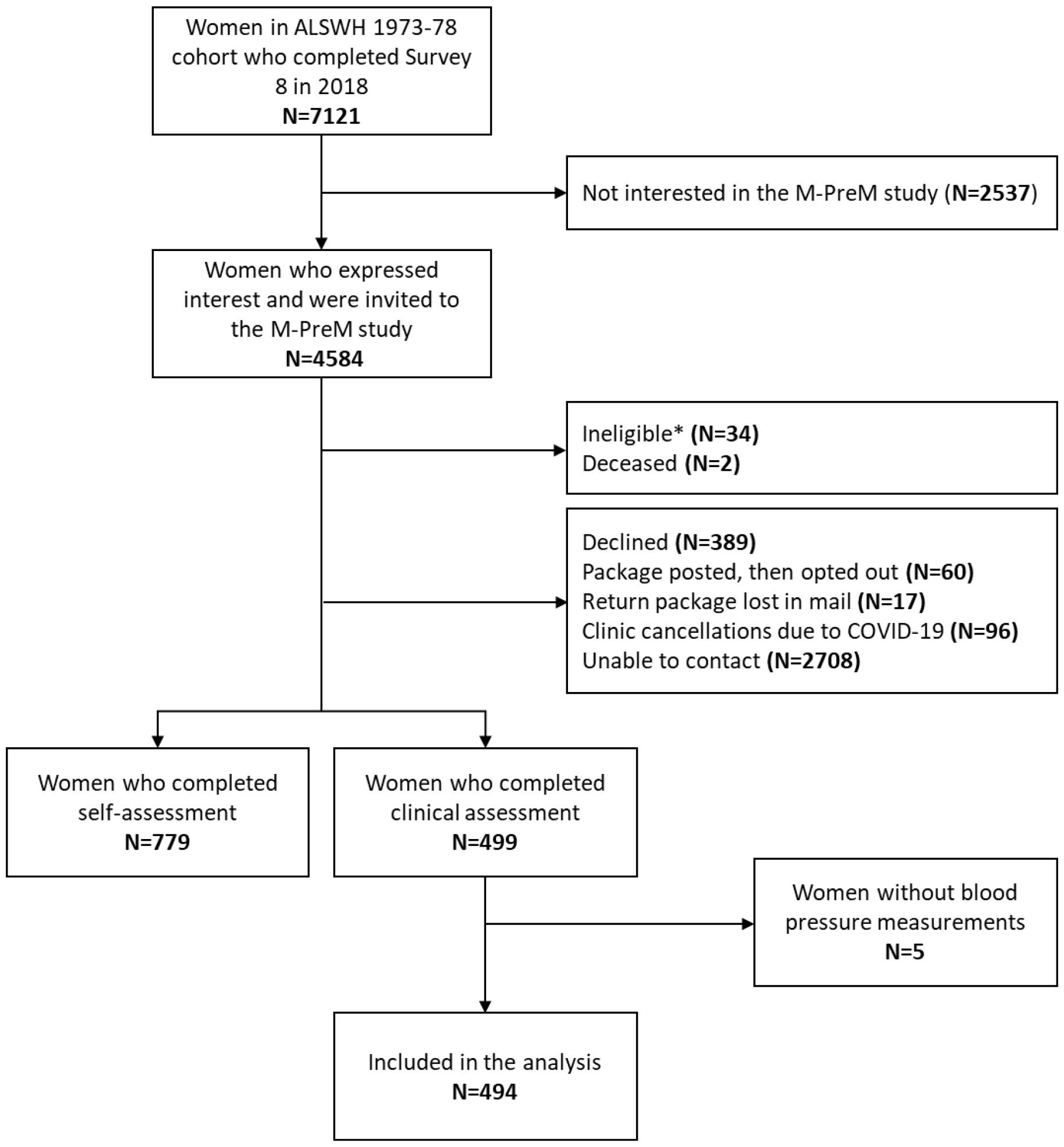
Flow chart of the selection of participants * Women who were pregnant or were being treated for a reproductive cancer at the time of invitation were excluded.

### Measurements of menstrual symptoms

We included three common menstrual symptoms in this study: heavy menstrual bleeding, irregular periods, and severe period pain. In each survey, questions regarding each menstrual symptom were asked: ‘*In the last 12 months, have you had heavy periods/irregular periods/severe period pain?*’ Women were defined as consistently having the symptom if their response was ‘often’, and as not consistently having the symptom if their response was ‘sometimes’, ‘rarely’ or ‘never’. Data collected in Survey 1 were not included since over-reporting of menstrual symptoms was found (also known as the ‘telescoping’ effect)^15^.

### Measurements of blood pressure

After subjects had rested in a seated position for 5 minutes, three measurements of SBP and DBP were recorded with an automatic blood pressure monitor, with a 1-2 minute interval between measurements. In accordance with the recommendations of the World Health Organization, the second and third measurements were averaged for analysis^19^.

### Covariates

Socio-demographic covariates included age (continuous), education (university degree or higher, trade/certificate/diploma, ≤12 years of schooling), and area of residence (classified as major cities, inner regional, and outer regional/remote based on an index of distance to the nearest urban centre^20^). Lifestyle factors included physical activity (measured as MET-minutes per week: nil/sedentary [0-39], low [40-599], moderate [600-1199], and high [≥1200])^21^ and smoking status (never, ex-smoker, and current smoker) ^22^.

Anthropometric measurements were collected in the M-PreM study. Weight without shoes was measured once using a digital scale, accurate to 0.1kg. Height without shoes was measured once using a stadiometer, accurate to 0.01m. Body mass index (BMI) was calculated by dividing weight in kilograms by the square of height in meters and classified as underweight/normal weight (<25kg/m^2^), overweight (25 to <30kg/m^2^), and obese (≥30kg/m^2^). The waist circumference was measured at the midpoint between the bottom of the last palpable rib and top of the iliac crest. The hip circumference was measured at the widest point of the buttocks. Waist-to-hip ratio was calculated.

### Statistical analysis

Means (±standard deviations) and numbers (percentages) were used to describe the characteristics of the study population. Latent class growth analysis (LCGA, with the ‘PROC TRAJ’ procedure in SAS^23^) was used to identify subgroups of participants following similar trajectories of each menstrual symptom from Survey 2 (2000) to Survey 8 (2018). The use of oral contraceptive pill (time-varying, dichotomized as ‘yes’ and ‘no’ in each survey) was included in the LCGA as the covariate to better separate the trajectories. The Bayesian information criterion was used to choose the best-fit model^24^. After determining the trajectories of each menstrual symptom, mean blood pressure was compared among different trajectories using the analysis of variance (ANOVA). The associations between trajectories of each menstrual symptom and blood pressure were assessed by linear regression models. Covariates in the models included age, education, area of residence, physical activity, smoking status collected in Survey 8, and BMI and waist-to-hip ratio collected in the M-PreM study. Since blood pressure was measured using different monitors, we also included blood pressure monitors as a covariate. All statistical analyses were performed using SAS 9.4, and a *P*-value < 0.05 was considered statistically significant.

## Results

The characteristics of the study population are shown in Table 1. The mean age of the subjects was 42.3 (±1.5) years in Survey 8. The majority lived in major cities (93.1%) and had a university degree or higher (74.9%), and more than half (59%) were overweight or obese. The proportion of women reporting often having heavy menstrual bleeding, irregular periods, and severe period pain in Survey 8 were 15.0%, 8.3%, and 6.1%, respectively.

**Table 1.** Characteristics of participants (N=494)

|  |  |
| --- | --- |
| Age, mean±SD | 42.3±1.5 |
| Area of residence, N (%) |  |
| Major cities | 458 (93.1) |
| Inner regional | 26 (5.3) |
| Outer regional/remote | 8 (1.6) |
| Education, N (%) |  |
| University degree or higher | 370 (74.9) |
| Trade/certificate/diploma | 90 (18.2) |
| ≤12 years of schooling | 34 (6.9) |
| Physical activity, N (%) |  |
| Nil/sedentary | 53 (11.0) |
| Low | 118 (24.6) |
| Moderate | 117 (24.4) |
| High | 192 (40.0) |
| Smoking status, N (%) |  |
| Never | 340 (68.8) |
| Ex-smoker | 124 (25.1) |
| Current smoker | 30 (6.1) |
| Current use of oral contraceptive pill, N (%) | 67 (13.6) |
| Menstrual symptoms |  |
| Heavy menstrual bleeding, N (%) | 74 (15.0) |
| Irregular periods, N (%) | 41 (8.3) |
| Severe period pain, N (%) | 30 (6.1) |
| **Measurements collected in M-PreM study** |  |
| Systolic blood pressure (mmHg), mean±SD | 119.1±12.4 |
| Diastolic blood pressure (mmHg), mean±SD | 77.3±8.8 |
| Body mass index, N (%) |  |
| Underweight/normal weight (<25kg/m<sup>2</sup>) | 202 (41.0) |
| Overweight (25 to <30kg/m<sup>2</sup>) | 146 (29.6) |
| Obese (≥30kg/m<sup>2</sup>) | 145 (29.4) |
| Waist-to-hip ratio, mean±SD | 0.8±0.1 |

As shown in Figure 2, LCGA identified the following group-based trajectories (see Supplemental Table 2A-2C for model selection): 3 classes for heavy menstrual bleeding (normative [79.4%], later onset [12.6%], and chronic [8.1%]), 2 classes for irregular periods (normative [88.7%] and later onset [11.3%]), and 2 classes for severe period pain (normative [84.9%] and chronic [15.1%]), where ‘normative’ represented consistently low or none symptoms, ‘chronic’ represented a high probability of menstrual symptoms in each survey, and ‘later onset’ represented an increasing probability in recent surveys.

**Figure 2.**
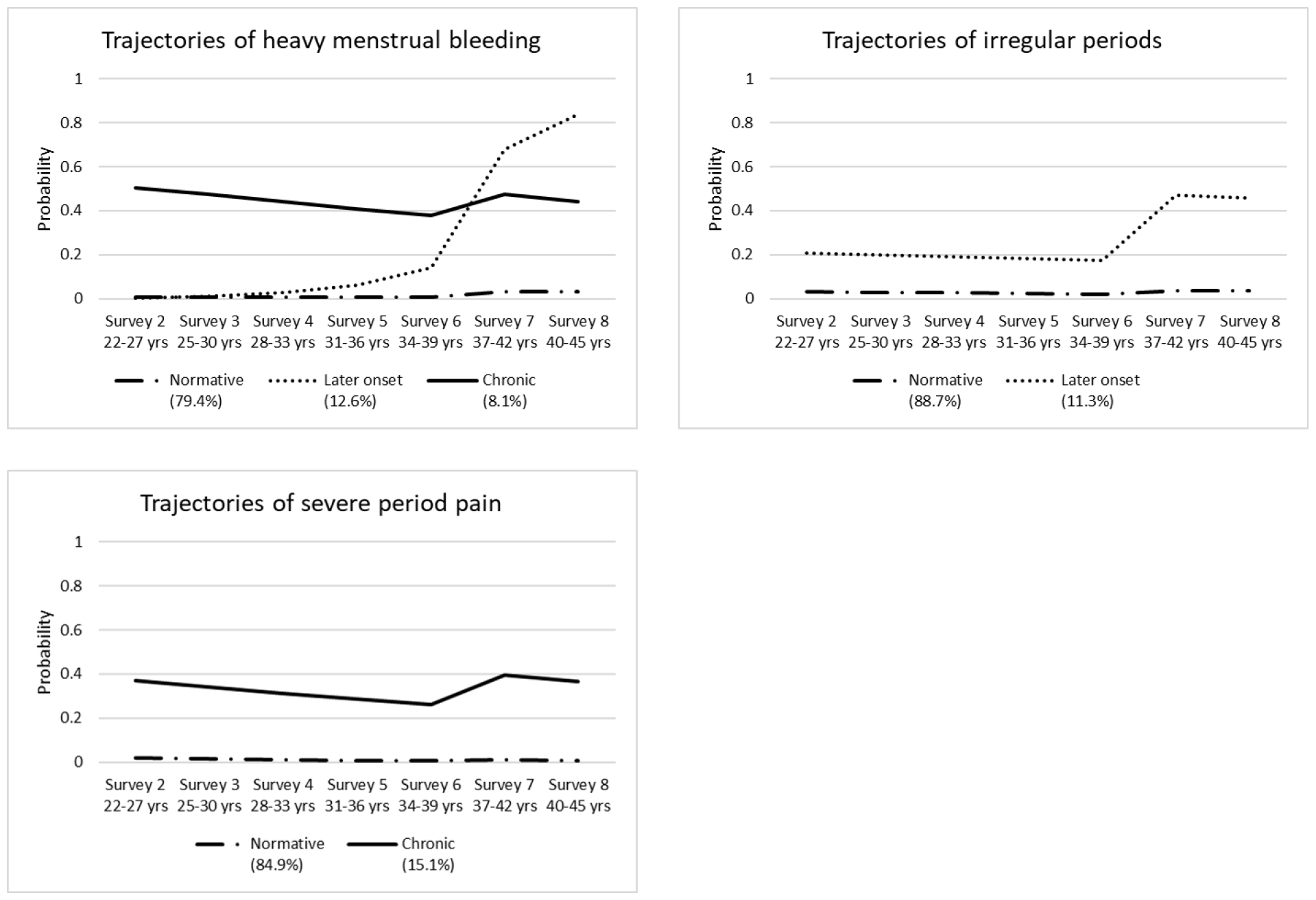
Trajectories of heavy menstrual bleeding, irregular periods, and severe period pain from Survey 2 (2000) to Survey 8 (2018), N=494

Table 2 shows the mean blood pressure by trajectories of heavy menstrual bleeding, irregular periods, and severe period pain. Women in different trajectories of heavy menstrual bleeding had different SBP and DBP. For trajectories of irregular periods, women in the later onset group had higher DBP than women in the normative group (80.1 versus 77.0 mmHg), and they were comparable regarding SBP. Women in different trajectories of severe period pain had similar SBP and DBP.

**Table 2.** Mean blood pressure by trajectories of heavy menstrual bleeding, irregular periods, and severe period pain.

|  | Systolic blood pressure |  | Diastolic blood pressure |  |
| --- | --- | --- | --- | --- |
|  | Mean±SD | P-value | Mean±SD | P-value |
| Trajectories of heavy menstrual bleeding |  |  |  |  |
| Normative (79.4%) | 118.3±12.0 | 0.01 | 76.5±8.6 | <0.001 |
| Late onset (12.6%) | 121.6±13.6 |  | 80.0±9.4 |  |
| Chronic (8.1%) | 123.9±12.7 |  | 81.3±8.9 |  |
| Trajectories of irregular periods |  |  |  |  |
| Normative (88.7%) | 119.0±12.5 | 0.5 | 77.0±8.9 | 0.02 |
| Late onset (11.3%) | 120.3±11.1 |  | 80.1±8.3 |  |
| Trajectories of severe period pain |  | 0.9 |  | 0.8 |
| Normative (84.9%) | 119.1±12.3 |  | 77.2±8.8 |  |
| Chronic (15.1%) | 119.0±13.2 |  | 77.6±8.9 |  |

**Table 3.** Associations of trajectories of heavy menstrual bleeding, irregular periods, and severe period pain with blood pressure.

|  | Model 1 | Model 2 | Model 3 |
| --- | --- | --- | --- |
| Systolic blood pressure |  |  |  |
| Trajectories of heavy menstrual bleeding |  |  |  |
| Normative (79.4%) | 0 (ref) | 0 (ref) | 0 (ref) |
| Late onset (12.6%) | 4.0 (0.6, 7.3) | 4.1 (0.8, 7.4) | 3.4 (0.3, 6.5) |
| Chronic (8.1%) | 6.0 (1.7, 10.2) | 5.9 (1.7, 10.2) | 3.0 (-1.1, 7.0) |
| Trajectories of irregular periods |  |  |  |
| Normative (88.7%) | 0 (ref) | 0 (ref) | 0 (ref) |
| Late onset (11.3%) | 1.4 (-2.4, 5.1) | 1.4 (-2.3, 5.1) | 0.9 (-2.6, 4.4) |
| Trajectories of severe period pain |  |  |  |
| Normative (84.9%) | 0 (ref) | 0 (ref) | 0 (ref) |
| Chronic (15.1%) | 0.3 (-2.9, 3.6) | 0.4 (-2.9, 3.6) | -0.5 (-3.6, 2.5) |
| Diastolic blood pressure |  |  |  |
| Trajectories of heavy menstrual bleeding |  |  |  |
| Normative (79.4%) | 0 (ref) | 0 (ref) | 0 (ref) |
| Late onset (12.6%) | 3.5 (1.3, 5.8) | 3.6 (1.4, 5.9) | 3.1 (1.0, 5.2) |
| Chronic (8.1%) | 4.8 (1.9, 7.6) | 4.8 (1.9, 7.7) | 2.5 (-0.3, 5.2) |
| Trajectories of irregular periods |  |  |  |
| Normative (88.7%) | 0 (ref) | 0 (ref) | 0 (ref) |
| Late onset (11.3%) | 2.8 (0.3, 5.3) | 2.7 (0.2, 5.3) | 2.3 (-0.1, 4.6) |
| Trajectories of severe period pain |  |  |  |
| Normative (84.9%) | 0 (ref) | 0 (ref) | 0 (ref) |
| Chronic (15.1%) | 0.5 (-1.7, 2.8) | 0.5 (-1.7, 2.8) | -0.2 (-2.3, 1.8) |

Model 1 was adjusted for blood pressure monitors.
Model 2 was further adjusted for age, education, and area of residence in Survey 8.
Model 3 was further adjusted for physical activity, smoking status in Survey 8 and body mass index and waist-to-hip ratio collected in M-PreM study.

After adjusting for blood pressure monitors and socio-demographic characteristics (age, education, and area of residence), for trajectories of heavy menstrual bleeding, women in the late onset and chronic group had higher SBP and DBP than women in the normative group (late onset group: B=4.1 [95% CI: 0.8, 7.4] for SBP, B=3.6 [1.4, 5.9] for DBP; chronic group: B=5.9 [1.7, 10.2] for SBP, B=4.8 [1.9, 7.7] for DBP). Further adjustment for lifestyle factors (physical activity and smoking status), BMI, and waist-to-hip ratio attenuated the estimates (late onset group: B=3.4 [0.3, 6.5] for SBP, B=3.1 [1.0, 5.2] for DBP and chronic group: B=3.0 [-1.1, 7.0] for SBP, B=2.5[-0,3, 5.2] for DBP). For trajectories of irregular periods, women in the late onset group had higher DBP than women in the normative group, after adjusting for blood pressure monitors and socio-demographic characteristics (B=2.7 [0.2, 5.3]). The estimate was attenuated after further adjusting for lifestyle factors, BMI, and waist-to-hip ratio (B=2.3 [-0.1, 4.6]). There was no difference between trajectories of irregular periods and SBP. We did not find associations between trajectories of severe period pain and blood pressure.

## Discussion

This study used longitudinal data collected over 18 years to examine the relationship between the trajectory of menstrual symptoms, including heavy menstrual bleeding, irregular periods, and severe period pain with changes to mean SBP and DBP. The findings from this study indicate that women with late onset heavy menstrual bleeding had higher SBP and DBP, while women with late onset irregular periods had higher DBP. Severe period pain was not associated with any changes to mean blood pressure.

### Strengths and Limitations

This study has several strengths. First, it uses 18 years of longitudinal data, with surveys conducted every 3 years, which enabled us to map changes in symptoms throughout the reproductive life span and thus build symptom trajectory profiles. Following on from this, we were able to use these symptom trajectories to delve deeper into the relationships between symptoms previously reported to be associated with blood pressure and determine how specific symptom trajectories are linked to mean SBP and DBP. Second, the prospectively collected information on menstrual symptoms and lifestyle variables were less likely to be prone to recall bias. Finally, we included a wide range of potential confounders in our statistical modelling, strengthening the confidence in the results of this study.

A limitation of the current study is that in recent years, the definition of heavy menstrual bleeding has been changed from a diagnosis based on estimates of menstrual blood loss to a more patient-centred approach. The review by the National Institute for Health and Care Excellence^25^ defines heavy menstrual bleeding as “any excessive menstrual blood loss which interferes with the woman’s physical, emotional, social, and material quality of life, and which can occur alone or in combination with other symptoms”. The question used in the ALSWH survey for heavy menstrual bleeding does not use identical criteria but does indicate the relationship between the symptoms experienced by women. Studies that have investigated the validity of women’s self-reported menstrual symptoms and these have primarily focussed on menstrual cycle length. One study, however, that quantified the volume of blood loss in heavy menstrual bleeding found an association with increased risk of hypertension ^26^. This supports the relationship found in the current study between reported heavy menstrual bleeding and higher measured blood pressure.

### Comparison with other studies

In our study, we found that the chronic heavy menstrual bleeding group and those with later onset heavy menstrual bleeding, from when participants were 31-36 years of age, were associated with increased SBP and DBP following adjustment for all covariates. We previously reported that young women who experienced heavy menstrual bleeding had an increased risk of incident chronic hypertension, compared with those who had not experienced heavy menstrual bleeding. This relationship is also likely bi-directional, with incident chronic hypertension increasing risk of heavy menstrual bleeding ^1^. The current study confirms previous reports of heavy menstrual bleeding increasing risk of high blood pressure, but also demonstrates that the trajectory of the symptom, specifically women who experience later onset heavy menstrual bleeding, are at greater risk of high blood pressure. Heavy menstrual bleeding is more likely to occur in older women as they approach menopause due to estrogen fluctuations, and increased risk of developing polyps or endometrial hyperplasia ^27^. Heavy menstrual bleeding has been reported to double in prevalence in women surveyed between the ages of 22 and 42 across a 15-year period, suggesting that a high proportion of women are likely to experience later onset heavy menstrual bleeding ^4^ and thus, are at an increased risk of high blood pressure. In our study, after adjusting for lifestyle behaviours (e.g., physical activity, smoking status, and BMI), the association between heavy menstrual bleeding and blood pressure was attenuated, indicating heathier lifestyles (e.g., increasing PA levels) may be helpful to alleviate the adverse impact of heavy menstrual bleeding on blood pressure.

A previous study reported increased risk of incident irregular periods due to chronic hypertension but, unlike heavy menstrual bleeding, the relationship did not appear to be bi-directional as irregular periods were not associated with increased risk of incident chronic hypertension ^1^. The current study further characterises the complexity of this relationship by demonstrating that women with later onset irregular periods, from the ages of 34-39 years, have a higher risk of an increased DBP following adjustment for all covariates. Menstrual cycle irregularities have been associated with high salivary testosterone levels in healthy women ^28^, with high testosterone seeming to increase renin activity and systolic blood pressure in women ^29^. In fact, chronically high testosterone induces several physiological changes that may contribute to the development of high blood pressure and promote development of cardiovascular disease ^30^. This suggests that women with irregular periods may have elevated testosterone levels which may be indirectly causing an increase in blood pressure. Future studies should determine testosterone levels in women with irregular periods.

The fact that no association was found between painful periods and blood pressure is not surprising, given that previous studies within this cohort have not reported such an association. However, the relationship between painful periods and blood pressure has been reported previously in several studies^26^. A 14-year follow up study reported that women with painful periods were also at increased risk of ischaemic heart disease if they are hypertensive ^31^, while a cohort of Japanese women had increased risk of developing hypertensive disorder of pregnancy if they had painful periods around the age of 20 ^32^, highlighting that the long term effects of painful periods on blood pressure should not be discounted.

Both heavy menstrual bleeding and irregular periods increased risk of high DBP. While the increase in DBP is only mild, one study demonstrated that a diastolic hypertension burden of at least 80mm Hg in the absence of significant changes to SBP, independently predicted risk of a composite outcome of myocardial infarction, ischemic stroke, or haemorrhagic stroke across a period of 8 years ^33^. Similarly, isolated diastolic hypertension has also been associated with higher risk of combined fatal with non-fatal cardiovascular and coronary events ^34^. Furthermore, in young participants (<50 years), DBP was the strongest predictor of coronary heart disease risk ^35^, total mortality, cardiovascular mortality and all cardiovascular (fatal and nonfatal) events ^34^. As all participants in the current study were less than 50 years of age, this means that women who present with symptoms of late onset heavy menstrual bleeding and irregular periods should be monitored for their DBP as they age.

Our results showed the trajectory of heavy menstrual bleeding and irregular periods are positively associated with blood pressure in midlife. As high blood pressure is a major risk factor for cardiovascular disease, including coronary heart disease, which is the second most common cause of death in Australian women ^36^, this study highlights that physicians should monitor their patients’ menstrual symptoms and blood pressure regularly, particularly women who develop late onset heavy menstrual bleeding or irregular periods. Future studies should investigate how these symptoms may be associated with blood pressure to increase their sensitivity as predictors of disease or to inform the development of preventative interventions.

## Data Availability

Data are available upon reasonable request. Access to the M-PreM dataset requires approval from the Australian Longitudinal Study on Women?s Health (ALSWH) Data Access Committee. More information can be found at the ALSWH website: https://alswh.org.au/for-data-users/.

## Acknowledgements

The authors would like to acknowledge the Australian Government’s Department of Health and Aged Care for funding the ALSWH. The authors would also like to thank the women who participated in ALSWH and the M-PreM study. The authors acknowledge Nykola Kent for her contribution to conducting the literature review and assisting in drafting the manuscript.

We acknowledge that the boundaries around what “women’s health” incorporates are not always clear, and that sex and gender are distinct concepts. Although it is necessary for us to collect information on health topics related to the female sex (i.e. menstruation, childbirth, menopause), we acknowledge that not everyone who identifies as a woman experiences these female-specific health issues or identifies as female.

## Funding

GDM is supported by the Australian National Health and Medical Research Council Leadership Fellow (APP2009577). CJ is supported by China Scholarship Council (grant number: 202006010042) (https://www.csc.edu.cn/chuguo/s/1844). The M-PreM Study is supported by the National Health and Medical Research Council Project Grant (APP1129592).

## Conflicts of interests

None

## Notes

### Competing Interest Statement

The authors have declared no competing interest.

### Clinical Trial

This study doesn't involve any interventions.

### Author Declarations

This study involves human participants and was approved by the Metro South Health and Health Services Human Research Ethics Committee (reference number: HREC/2019/QMS/52052) and ratified by the University of Newcastle and the University of Queensland Human Research Ethics Committees. All participants provided informed consent by completing an electronic or paper participant consent form.

